# Surveillance patterns in non-muscle invasive bladder cancer across risk groups: a real-world analysis

**DOI:** 10.1101/2025.11.25.25340944

**Authors:** Lisa M.C. van Hoogstraten, Sita H. Vermeulen, Jasper P. Hof, Antoine G. van der Heijden, Alina Vrieling, Lambertus A. Kiemeney, Katja K.H. Aben

**Affiliations:** Department of Research and Development, Netherlands Comprehensive Cancer Organisation, Utrecht, The Netherlands; Department of Urology, Radboud University Medical Center, Nijmegen, The Netherlands; IQ Health Science Department, Radboud University Medical Center, Nijmegen, The Netherlands; Center for Quantitative Genetics and Genomics, Aarhus University, Aarhus, Denmark

**Keywords:** Non-muscle invasive bladder cancer, surveillance, real-world data, practice patterns, guideline adherence, bladder cancer

## Abstract

**OBJECTIVE:** To provide insight into real-world surveillance practices for patients with low risk (LR), intermediate risk (IR), and (very) high risk (HR) non-muscle invasive bladder cancer (NMIBC).

**MATERIALS AND METHODS:** Cystoscopy surveillance patterns were analysed using real-world data from two population-based cohort studies in the Netherlands, comparing practices to guideline recommendations by risk group. As bladder cancer-related symptoms may prompt diagnostic investigations beyond routine follow-up, we evaluated the occurrence of such symptoms and the use of various diagnostics including cystoscopy in a subset of patients with available data.

**RESULTS:** In total, 2,791 primary and recurrent tumours were included in the analyses. Among patients with LR NMIBC, 37.6% were monitored more intensively than recommended. The average number of cystoscopies in the first follow-up year was 1.3 (range 0-4). IR NMIBCs were generally monitored in accordance with guideline recommendations. However, adherence declined over the years following diagnosis, dropping from 78.1% to 59.3%. The proportion of IR NMIBCs who were monitored less than recommended increased from 21.7% to 39.8%. Most HR NMIBCs (88.2%) were monitored less than recommended, though adherence improved over time. In-depth analysis of 204 tumours revealed that next to cystoscopy, cytology was frequently employed, increasing with risk group (LR: 50.0%, IR: 52.3%, HR: 88.9%). Imaging (25%-65%) and biopsies (25%-85%) were also commonly performed. Surveillance patterns varied especially among IR NMIBCs. Symptoms were reported in approximately one third of bladder tumours during follow-up, but did not appear to affect surveillance patterns.

**CONCLUSION:** Our findings demonstrate substantial deviations from recommended NMIBC surveillance practices. Cystoscopy was overused in a considerable proportion of LR NMIBC and underused in most HR cases. Surveillance of IR NMIBCs was particularly heterogenous with considerable variation in diagnostic investigations. These insights highlight opportunities to refine NMIBC surveillance schedules, aiming to improve patient outcomes and optimize allocation of healthcare resources.

## INTRODUCTION

The vast majority of patients with bladder cancer are diagnosed with non-muscle invasive bladder cancer (NMIBC). The prognosis of NMIBC is generally good, however, depending on tumour stage and grade, many patients experience disease recurrence despite treatment(1). There is also a probability of progression to muscle-invasion and metastatic disease, which is associated with significantly worse outcomes(2, 3). Due to the risk of recurrence and progression, international guidelines recommend frequent monitoring.

The European Association of Urology (EAU) has formulated recommendations for follow-up, based on the stratification of patients into risk groups(4). Based on their probability of tumour progression(5), patients are categorized into low risk (LR), intermediate risk (IR), and (very) high risk (HR) NMIBC. Although the guidelines and definition of the risk groups have changed over time, the concept remained the same: a LR bladder tumour necessitates less intensive monitoring and treatment compared to a HR bladder tumour.

Although urinary markers are increasingly used, cystoscopy still remains the cornerstone of NMIBC surveillance. The EAU guidelines recommend the first cystoscopy after transurethral resection of the bladder tumour (TURBT) at 3 months in all risk groups, as it serves as a key prognostic indicator for both disease recurrence and progression(4). An increase in risk necessitates a corresponding escalation on both the intensity and duration of surveillance (**Figure 1, Table 1**). For patients in the (very) high risk group, surveillance with cystoscopy should be combined with cytology and CT urography.

**Table 1:** Guideline recommendations regarding follow-up intensity for NMIBC, stratified per risk group.

| Risk group | EAU (2016) |  |  |  | EAU (2024) |  |  |  |
| --- | --- | --- | --- | --- | --- | --- | --- | --- |
|  | Cytology | Cystoscopy | Imaging | Duration of follow-up | Cytology | Cystoscopy | Imaging | Duration of follow-up |
| Low risk | No | At 3 and 12 months, then annually | Not systematic | 5 years | No | At 3 and 12 months, then annually | Not systematic | 5 years |
| Intermediate risk | Not systematic | At 3 months, then in between low- and high risk schedule | Not systematic | In between | No | At 3 months, every 6 months for 2 years, then annually for 10 years | Not systematic | 10 years |
| High/very high risk | Yes | Every 3 months for 2 years, every 6 months up to 5 years, then annually | Yearly | Life long | Yes | Every 3 months for 2 years, every 6 months up to 5 years, then annually | Annually up to 5 years Then every 2 years up to 10 years | Life long |

NMIBC: non-muscle invasive bladder cancer; EAU: European association of Urology

**Figure 1:**
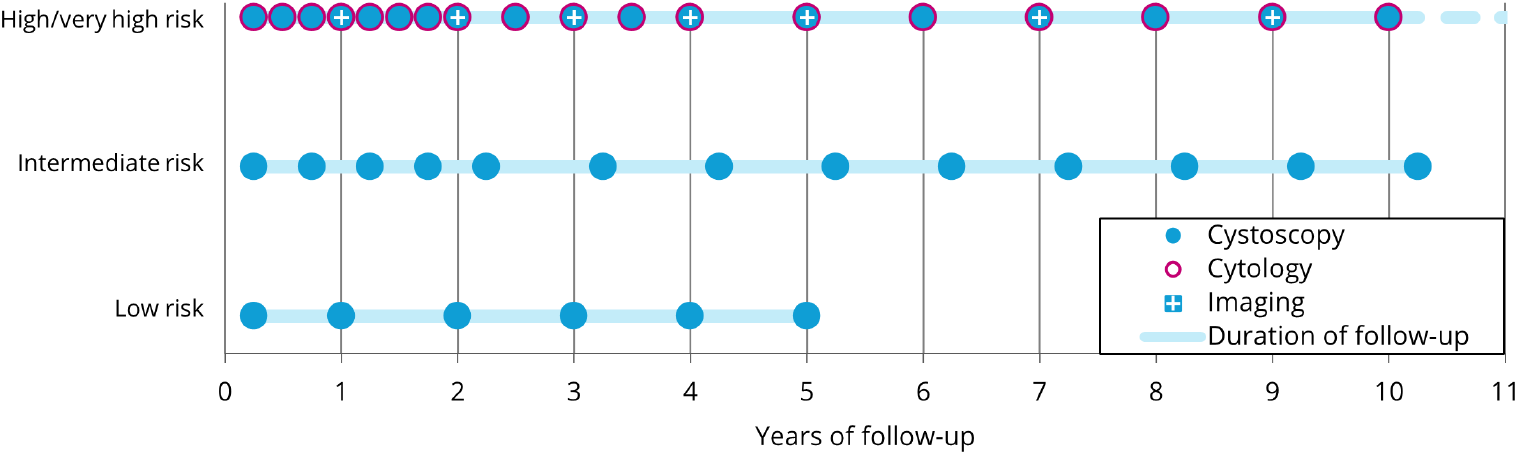
Follow-up protocol for NMIBC according to the EAU 2024 guidelines by risk group. NMIBC: non-muscle invasive bladder cancer

Surveillance schedules for NMIBC, both current and historical, are not supported by high level evidence(4, 6). Instead, the recommendations are largely based on expert opinion, a collaborative review(7) and a single clinical trial published in 1995 including less than 100 patients with IR NMIBC(8). As a result, the optimal frequency and duration of surveillance remain uncertain, and practice variation is expected. Previous research, although suffering from methodological shortcomings, indicated substantial overuse of cystoscopies and variation in cystoscopy intensity in patients with NMIBC(9–12). One potential explanation for these findings is that further diagnostic testing may be driven by bladder cancer-related symptoms experienced by patients. Such deviations from the guidelines may reflect personalized care rather than unwanted variation. However, these studies found that increased surveillance intensity did not lead to higher recurrence detection rates.

Bladder cancer is one of the most expensive cancer types per patient(13) and healthcare systems are under enormous pressure(14). Optimized evidence-based surveillance strategies are essential in order to make the most efficient use of healthcare resources without compromising patient outcomes. For this purpose, it is important to obtain insight into practice patterns. However, comprehensive real-world evidence on long-term surveillance practices and adherence to guideline-based surveillance in NMIBC is largely lacking. Therefore, the objective of this study was to provide contemporary data on surveillance practices in the Netherlands for NMIBC risk groups.

## MATERIALS AND METHODS

### Patient population

For this study, we used patient data from two population-based cohort studies performed in the South-Eastern part of the Netherlands; the UroLife study and Nijmegen Bladder Cancer Study (NBCS). Data collection for both studies was embedded in the Netherlands Cancer registry (NCR), maintained by the Netherlands Comprehensive Cancer Organisation (IKNL). Detailed descriptions of both cohorts have been published previously(15–17). For the current study, we included patients who were diagnosed between 2011-2021.

### Data collection

Data were available regarding patient and tumour characteristics (age, sex, smoking status, tumour stage, grade, carcinoma in situ (CIS), focality), treatment (TURBT, bladder instillations with chemotherapy or BCG, cystectomy) and cystoscopy surveillance. To evaluate surveillance patterns beyond the use of cystoscopy, we conducted an additional manual data collection from the electronic health records of a subset of patients to assess the use of cytology, imaging, biopsy, and biomarkers, with a minimum follow-up of 5 years. Since patient-reported symptoms may lead to advanced or additional diagnostic investigations, which may explain the observed follow-up beyond regular appointments, we also collected data on the occurrence of haematuria and other bladder cancer-related symptoms (e.g., urgency, frequency, pain) potentially indicating recurrence of the disease. We randomly selected patients with low, intermediate and (very) high risk NMIBC (Ta, Tis, T1) diagnosed in 4 different hospitals, including one university hospital and three non-academic teaching hospitals that participated in both cohort studies. Because of feasibility, this additional data collection was performed in a subset of 159 patients.

### Definitions

All primary and recurrent NMIBCs were categorized into low, intermediate or (very) high risk using an adapted version of the 2016 EAU risk group classification(18) due to incomplete data in the electronic health records, particularly regarding tumour size. The adapted EAU risk groups are based on tumour stage and grade; low risk: a primary, solitary, Ta low grade (LG) tumour. The very high risk group was not yet introduced in 2016, therefore the high and very high risk group were combined and include at least one of the following features: stage T1, high grade, CIS, or multiple recurrent Ta LG tumours. Intermediate risk: all tumours not defined in the low- and high risk category.

A recurrence was only considered after a tumour-free status had been achieved. For stage Ta/T1 tumours, a tumour-free status was reached after undergoing radical TURBT or re-TURBT, or in case of nonradical (re-)TURBT, after tumour-negative cystoscopy. Patients with stage Tis or concomitant CIS were considered tumour-free after a tumour-negative cystoscopy and, if applicable, negative cytology. A negative result was also required, if a biopsy was performed. Recurrence was defined as either a bladder tumour histologically confirmed by TURBT, fulguration ≥6 months after the previous TURBT (coded as TaG1 recurrence), a bladder tumour histologically confirmed after cystectomy, or progression to muscle-invasive bladder cancer (MIBC) or metastatic disease (mBC) after an initial NMIBC diagnosis.

Progression was defined as advancement to a higher disease stage, progression to MIBC (stage T2 or higher), regional lymph node involvement (N+), or distant metastasis (M+), as well as progression from low grade to high grade disease. Additionally, progression included radical cystectomy due to therapy resistance.

After each recurrence or progression, the NMIBC risk group was re-evaluated to allow for a correct comparison with the guideline-recommended surveillance protocol. For example, a patient initially diagnosed with Ta LG (LR NMIBC) who then experiences a Ta LG recurrence would be reassigned to the IR group, with the corresponding surveillance schedule reapplied from that point in time. If the disease subsequently progresses to T1 (HR NMIBC), the patient moves to the HR group and the HR surveillance protocol would be initiated.

Start of follow-up was defined as the first surveillance cystoscopy after TURBT and re-TURBT (if applicable). End of follow-up was defined as the last clinical contact between patient and urologist, radical cystectomy, progression to MIBC or mBC, or another reason for end of follow-up (i.e., death, moving abroad, diagnosis of other tumour with metastasis, other, unknown), whichever occurred first.

### Analysis

All analyses were performed at tumour level and per risk group. Descriptive analyses were performed to provide insight into the patient, tumour and treatment characteristics, per risk group and in total. Cystoscopy patterns were evaluated by calculating the average number of cystoscopies for each consecutive year after the first surveillance cystoscopy, provided that at least one cystoscopy was performed, and a complete year of follow-up was available. These averages were compared to the recommended annual number of cystoscopies per follow-up year according to the 2016 EAU guidelines and categorized into monitoring less than recommended, as recommended, and more than recommended.

In addition, the median time interval between consecutive cystoscopies was compared with the guideline-recommended time intervals if at least 2 cystoscopies were performed. As the 2016 guidelines do not specify the number of cystoscopies or time intervals for IR NMIBC, we also evaluated our data on IR NMIBC in light of the 2024 guidelines(18, 19). For the subset of patients for which more in-depth data were collected, individual surveillance trajectories following the first surveillance cystoscopy were visually evaluated through swimmer plots. All analyses were performed using SAS version 9.4 (SAS Institute, Cary, North Carolina, USA). Graphs were created using Excel.

Both the UroLife study (CMO 2013-494) and NBCS (CMO 2005-315) were approved by an accredited ethics committee in the Netherlands.

## RESULTS

In total, 2,791 primary and recurrent tumours from 1,890 unique patients were included in the study. Baseline characteristics of all NMIBCs are presented in **Table 2**.

**Table 2:** Patient, tumour and treatment characteristics of all NMIBCs (primary and recurrent), in total and by risk group.

|  | All NMIBCs<br>(N=2791) |  | Low Risk<br>(N=431) |  | Intermediate Risk<br>(N=1035) |  | High/very high<br>Risk (N=1325) |  |
| --- | --- | --- | --- | --- | --- | --- | --- | --- |
|  | N | (%) | N | (%) | N | (%) | N | (%) |
| **Patient** |  |  |  |  |  |  |  |  |
| **Age at diagnosis** (median, IQR) | 68.0 | (61.0-73.0) | 65.0 | (58.0-71.0) | 67.0 | (61.0-73.0) | 68.0 | (63.0-74.0) |
| **Gender** |  |  |  |  |  |  |  |  |
| Female | 605 | (21.7%) | 112 | (26.0%) | 254 | (24.5%) | 239 | (18.0%) |
| Male | 2186 | (78.3%) | 319 | (74.0%) | 781 | (75.5%) | 1086 | (82.0%) |
| **Smoking status** |  |  |  |  |  |  |  |  |
| Never smoker | 426 | (15.3%) | 63 | (14.6%) | 165 | (15.9%) | 198 | (14.9%) |
| Former smoker | 1704 | (61.1%) | 254 | (58.9%) | 616 | (59.5%) | 834 | (62.9%) |
| Current smoker | 611 | (21.9%) | 107 | (24.8%) | 240 | (23.2%) | 264 | (19.9%) |
| Unknown | 50 | (1.8%) | 7 | (1.6%) | 14 | (1.4%) | 29 | (2.2%) |
| **Hospital of diagnosis** |  |  |  |  |  |  |  |  |
| General hospital | 720 | (25.8%) | 133 | (30.9%) | 243 | (23.5%) | 344 | (26.0%) |
| Non-university teaching hospital | 1958 | (70.2%) | 274 | (63.6%) | 758 | (73.2%) | 926 | (69.9%) |
| University hospital | 113 | (4.0%) | 24 | (5.6%) | 34 | (3.3%) | 55 | (4.2%) |
| **Tumour** |  |  |  |  |  |  |  |  |
| **Clinical tumour stage** |  |  |  |  |  |  |  |  |
| Ta | 1807 | (64.7%) | 431 | (100.0%) | 1035 | (100.0%) | 341 | (25.7%) |
| T1 | 641 | (23.0%) | - |  | - |  | 641 | (48.4%) |
| CIS (Tai, Tis, T1i) | 343 | (12.3%) | - |  | - |  | 343 | (25.9%) |
| **Differentiation grade** |  |  |  |  |  |  |  |  |
| Low grade (G1, G2, G2A) | 1635 | (58.6%) | 431 | (100.0%) | 1031 | (99.6%) | 173 | (13.1%) |
| High grade (G2B, G3) | 1142 | (40.9%) | - |  | - |  | 1142 | (86.2%) |
| Unknown | 14 | (0.5%) | - |  | 4 | (0.4%) | 10 | (0.8%) |
| **Focality** |  |  |  |  |  |  |  |  |
| Unifocal | 1735 | (62.2%) | 431 | (100.0%) | 578 | (55.8%) | 726 | (54.8%) |
| Multifocal | 1023 | (36.7%) | - |  | 457 | (44.2%) | 566 | (42.7%) |
| Unknown | 33 | (1.2%) | - |  | - |  | 33 | (2.5%) |
| **Primary or recurrent tumour** |  |  |  |  |  |  |  |  |
| Primary | 1890 | (67.7%) | 431 | (100.0%) | 444 | (42.9%) | 1015 | (76.6%) |
| Recurrent | 901 | (32.3%) | - |  | 591 | (57.1%) | 310 | (23.4%) |
| **Treatment** |  |  |  |  |  |  |  |  |
| **reTURBT** | 778 | (27.9%) | 22 | (5.1%) | 61 | (5.9%) | 695 | (52.5%) |
| **Chemotherapy instillations\*** | 1659 | (59.4%) | 327 | (75.9%) | 670 | (64.7%) | 662 | (50.0%) |
| **BCG instillations\*** | 973 | (34.9%) | 0 | (0.0%) | 68 | (6.6%) | 905 | (68.3%) |

NMIBC: non-muscle invasive bladder cancer; IQR: interquartile range; TURBT: transurethral resection of the bladder tumour; BCG: Bacillus Calmette-Guérin
\*Treated at some point with chemotherapy/BCG instillations

### Cystoscopy patterns

In **Figure 2**, the adherence to the guideline recommendations per consecutive follow-up year are presented by risk group. Furthermore, median time intervals between consecutive cystoscopies are displayed in **Figure 3**. In **Suppl. Table 1** and **Suppl. Table 2**, more detailed data underlying the figures are depicted.

**Figure 2:**
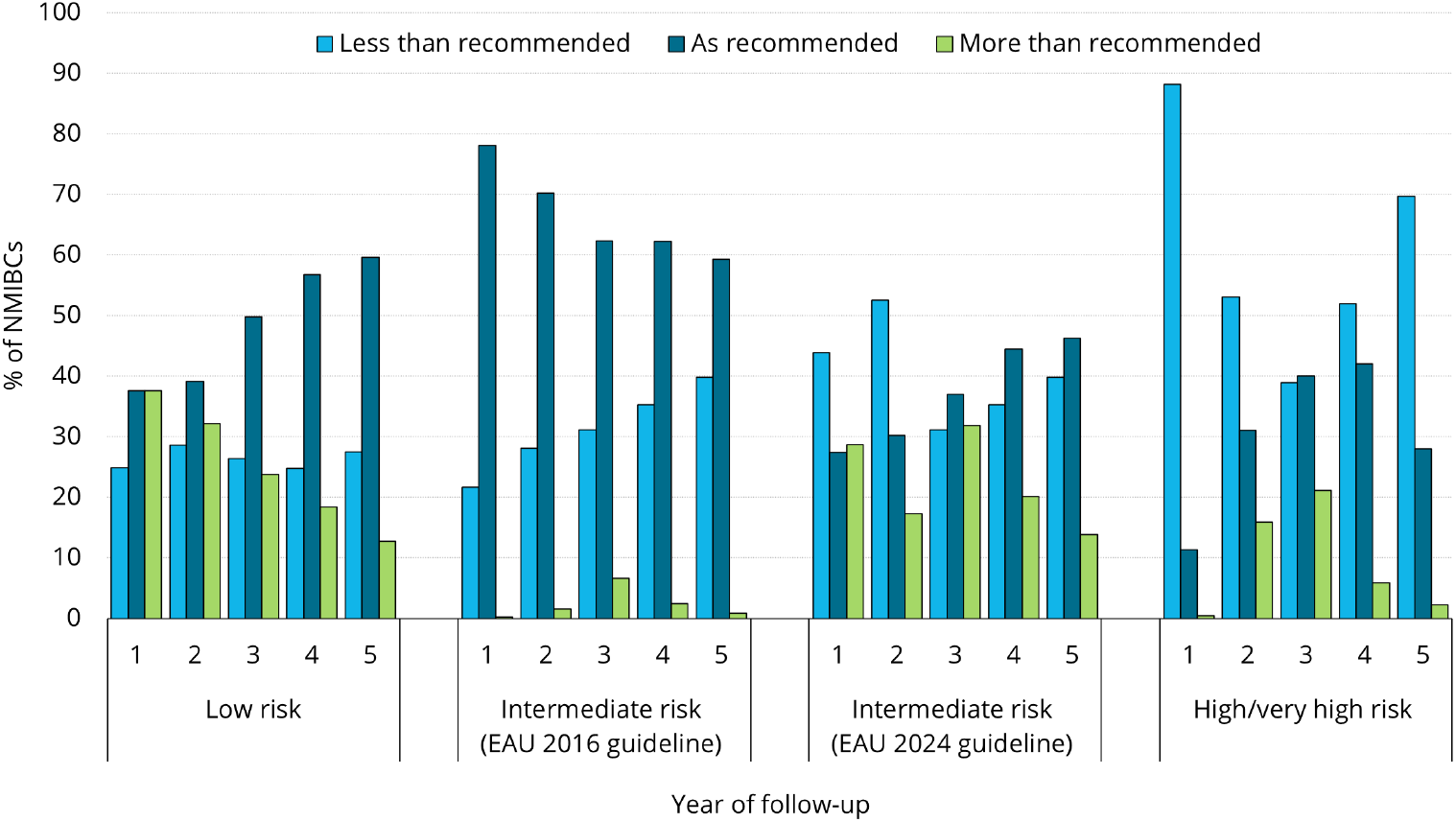
Monitoring of NMIBC per year of follow-up according to EAU recommendations*, stratified by risk group. *The recommended number of cystoscopies was calculated <u>after</u> the first surveillance cystoscopy. Low risk: 1 cystoscopy yearly. Intermediate risk (EAU 2016): 1-4 cystoscopies in the first year, 1-3 in the second year, and thereafter 1-2 cystoscopies yearly. Intermediate risk (EAU 2024): 2 cystoscopies in year 1 and 2, and 1 cystoscopy in subsequent years. High risk: 4 cystoscopies in the first year, 3 cystoscopies in the second year and thereafter 2 cystoscopies yearly. NMIBC: non-muscle invasive bladder cancer; EAU: European Association of Urology

**Figure 3:**
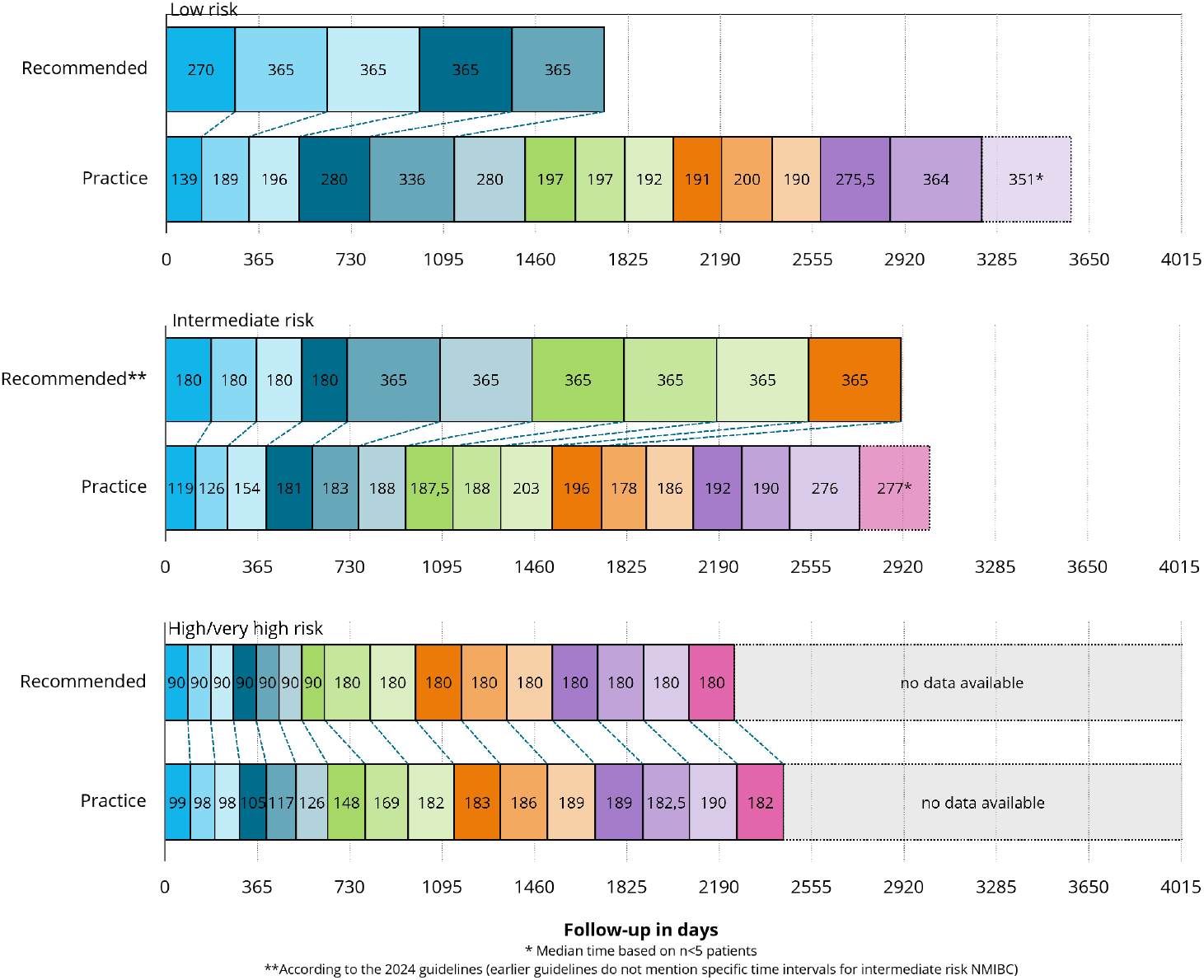
Median time interval (in days) between consecutive cystoscopies as recommended by the EAU NMIBC guidelines and in practice, stratified by risk group. NMIBC: non-muscle invasive bladder cancer; EAU: European Association of Urology

For LR NMIBC, the average number of cystoscopies in the first year of follow-up was 1.3 (SD ± 1.1, range 0-4). In half of the cases, the number of cystoscopies performed in real-world practice exceeds the recommended frequency. Some patients underwent more than twice the recommended number of cystoscopies (i.e., ≥10), with follow-up extending beyond five years of follow-up. The proportion of LR NMIBCs monitored according to guideline recommendations increased during follow-up from 37.6% in the first year to 59.6% in the fifth year. In contrast, 37.6% of LR NMIBCs were initially monitored more intensively than recommended, receiving up to 3 additional cystoscopies beyond the first surveillance cystoscopy. By the fifth year of follow-up, this proportion had decreased to 12.8%. As expected given the observed high surveillance intensity, the median interval between consecutive cystoscopies was shorter than recommended. Over the years following diagnosis, this interval was consistently below the advised 365 days, ranging from 29 to 177 days shorter, indicating surveillance intensity approaching nearly twice the recommended frequency.

For IR NMIBC, on average 1.7 (SD ± 1.2, range 0-5) cystoscopies were performed in the first year of follow-up. IR NMIBCs were generally monitored in line with the 2016 guideline recommendations. However, adherence decreased from 78.1% to 59.3% by the fifth year of follow-up. Conversely, the proportion of IR NMIBCs monitored less intensively than recommended, i.e., receiving no cystoscopy after the first follow-up cystoscopy at approximately 3 months, increased from 21.7% to 39.8%. This variation was expected, as the 2016 guidelines consider a broad range of cystoscopies as recommended. In caser the more specific 2024 guidelines were considered, no clear pattern emerged; most IR NMIBCs appeared to be monitored as recommended or less than recommended, although 28.7% of IR NMIBCs received up to 3 additional cystoscopies in the first year of follow-up, but this proportion declined by year five to 13.9%. The intervals between consecutive cystoscopies were generally shorter than stated in the guidelines, particularly after the transition from biannual to annual monitoring. The difference in timing ranged from 162 to 182 days below the advised interval).

For HR NMIBC, an average of 2.3 (SD ± 1.2, range 0-5) cystoscopies was performed in the first year of follow-up. The majority of HR NMIBCs (88.2%) were monitored less than recommended in the first year of follow-up, i.e. receiving less than 4 or even no cystoscopies, besides the first follow-up cystoscopy at approximately 3 months. During follow-up, this proportion decreased, and more HR NMIBCs received the recommended number of cystoscopies. For HR NMIBC, the average time interval was longer than recommended, mainly in the first two years of follow-up (difference range: 8-58 days). After the transition from every 3 months to biannual follow-up, median time intervals were similar to the recommended interval (ranging from 11 days below or 10 days above).

### Surveillance patterns beyond cystoscopy

In-depth surveillance patterns were evaluated in 204 primary and recurrent bladder tumours from 159 patients; 44 LR (21.6%), 88 IR (43.1%), and 72 HR (35.3%) NMIBC (**Suppl. Table 3**). In the vast majority (n=199, 97.5%) at least the first surveillance cystoscopy following diagnosis of the primary bladder cancer was performed. Additionally, 75.0% (n=153) of cases received one or more subsequent cystoscopies. Among the 25% (n=51) of cases that did not undergo further cystoscopic surveillance, 36 (70.6%) received alternative follow-up such as cytology, biopsy or CT urography/IVP. For the remaining cases, specifically 6 LR, 7 IR and 2 HR tumors, no further follow-up investigations were documented.

For the 44 primary LR NMIBCs, follow-up after the first surveillance cystoscopy consisted mainly of cystoscopies at fairly regular intervals, with cytology performed in 50.0% (N=22) of cases (**Suppl. Figure 1**). Imaging and biopsies were each used in 25.0% (N=11).

Among the 88 IR NMIBCs, surveillance strategies varied considerably in terms of type of diagnostics used, frequency and duration (**Suppl. Figure 2**). Cystoscopy was the main diagnostic tool (96.6%, N=85), though with varying intervals. Some patients followed a low-risk-like schedule with annual cystoscopies, while others received more intense follow-up resembling high-risk protocols. Cytology (52.3%, N=46), imaging (28.4%, N=25) and biopsy (36.4%, N=32) were also frequently employed.

Surveillance of 72 HR NMIBCs (**Suppl. Figure 3**) predominantly involved cystoscopy (97.2%, N=70) and cytology (88.9%, N=64), often performed at the same time. Biopsies were employed in 77.8% (N=56) of the cases, imaging in 55.6% (N=40). Patients with (concomitant) CIS were more likely to undergo cytology (95.0% vs. 86.5%), biopsy (85.0% vs. 75.0%) and imaging (65.0% vs. 51.9%) compared to patients without CIS (*data not shown*). Use of biomarkers, i.e., the Bladder EpiCheck, was rare, applied in only one HR and two IR cases, and was therefore not further evaluated. This was expected as biomarkers were not widely applied yet in the period of the study cohort.

Symptoms were reported in 45.5% (N=20) of LR NMIBC, 26.1% (N=23) of IR NMIBC, and 34.7% (N=25) of HR NMIBC. As shown in the swimmer plots (**Suppl. Figure 1-3**), most symptoms were followed by- or coincided with diagnostic investigations, mainly cystoscopy, as recommended by guidelines. The presence of symptoms did not appear to affect the surveillance pattern; the timing and type of follow-up investigations remained consistent with pre-symptom trajectories across all risk groups.

## DISCUSSION

In this study we evaluated surveillance patterns in daily clinical practice for patients with NMIBC per risk group. During the first year of follow-up, over one-third of patients with LR-NMIBNC were monitored more frequently and logically, time intervals between consecutive cystoscopies were shorter than recommended by the guidelines. In IR NMIBC, this proportion was over one quarter. The overuse of cystoscopy for both LR NMIBC and IR NMIBC decreased in subsequent years of follow-up. A different pattern was observed in HR NMIBC as the vast majority were monitored less intensively and in the first two year of follow-up time intervals between consecutive cystoscopies were slightly shorter than recommended. Next to cystoscopy, cytology, imaging and biopsy were also regularly performed during follow-up, with their use increasing across higher risk groups. The type of diagnostic procedures, as well as the frequency and duration of surveillance, varied between cases, especially within the IR NMIBC. Although symptoms were frequently reported in IR and HR NMIBC cases, their presence did not appear to impact the surveillance strategy.

Real-world, long-term surveillance of NMIBC is an understudied topic and current practice patterns are largely unknown. Evidence supporting existing guidelines is limited(7, 8), and much of the literature on practice variation suffers from methodological shortcomings, such as reliance on arbitrary cut-offs to define surveillance intensity, only evaluating cystoscopy surveillance practices, and short follow-up durations, often with a maximum of two years. Nevertheless, our findings are in line with the best available evidence(9-11, 20, 21). The phenomenon of surveillance overuse in low risk patients and underuse in high risk patients has been documented in other cancer types, such as colorectal cancer(22), breast cancer(23), and prostate cancer(23, 24). Until now, explanations for these findings remain unclear. The observed suboptimal guideline adherence in the type, frequency and duration of surveillance imply that risk-adapted, guideline-recommended surveillance is not widely implemented in clinical practice.

A possible explanation for the suboptimal guideline adherence could be that surveillance may be driven by the presence of symptoms. However, no clear differences in guideline adherence were observed in patients experiencing symptoms and the entire study population. Symptoms were frequently reported, and typically followed by diagnostic investigations, yet these did not lead to noticeable adjustments in the surveillance strategy. Other explanations for the observed practice variation include the limited evidence supporting current guideline recommendations(25), insufficient awareness or understanding of the guidelines and risk stratification(26), patient preferences(27), and logistical challenges. These challenges encompass scheduling issues, resource availability, and delays related to or resulting from treatment(28).

Variation in surveillance was particularly pronounced among patients with IR NMIBC, especially in terms of the type of diagnostic investigations, timing and duration of surveillance. EAU guidelines consider IR NMIBC as an in-between risk group which should be monitored as such, but until recently, no specific recommendations were formulated. In 2024, recommendations became more explicit by stating specific cystoscopy intervals(19), but again, evidence supporting these recommendations is lacking. Given the heterogeneity within IR NMIBC, the current one-size-fits-all approach may be inadequate. Emerging literature suggests that IR NMIBC should be further sub-stratified into subgroups with corresponding surveillance intensity(29, 30). In our study, some surveillance trajectories seemed to resemble more of a LR NMIBC follow-up strategy, i.e., cystoscopies at yearly intervals, and others seemed to follow a HR NMIBC strategy, including cytology and imaging as well. We did not define explicit subgroups for further stratification of IR NMIBC, as this was outside the scope of the study. For now, surveillance for the intermediate risk group remains an unresolved challenge and further research is warranted to identify optimal, tailored surveillance strategies for this diverse patient population.

This study is, to our knowledge, the first to evaluate long-term surveillance patterns of NMIBC in real-world practice. By using population-based, high-quality data from two large cohorts, reflection of clinical practice was pursued. The additional manual data collection allowed for an evaluation of surveillance beyond cystoscopy use, providing relevant insights and directions for future research to improve clinical care. Nevertheless, some limitations should be acknowledged. First, due to insufficient availability of data on tumour size in the electronic patient records, we applied an adapted risk classification. However, among patients with available tumour size data, only 3% were reclassified to a lower risk group when using the original EAU classification, suggesting minimal impact on overall findings. Second, although the UroLife and NBCS study reflect care in a large part of the Netherlands, i.e. South-Eastern part, and likely reflect national practice, some degree of patient selection cannot be excluded, potentially limiting generalizability. The NBCS cohort consists of patients that were invited in batches over different time periods, using both incident and prevalent sampling. Prevalent sampling may introduce selection bias, since patients had to survive and still have NMIBC at time of inclusion. However, we expect its impact to be limited as the prognosis NMIBC is generally good. In addition, this method also allowed for the creation of a large, diverse cohort with extended follow-up. Finally, manual data collection on additional diagnostic investigations and symptoms was limited to a subset of the cohort. Preferably, these data would have been available for the entire cohort but this was not feasible. Nonetheless, the subset was randomly selected across multiple hospitals, ensuring representativeness of the broader cohort.

## CONCLUSION

This study highlights significant variation in surveillance practices for non-muscle invasive bladder cancer, indicating a gap between guideline recommendations and real-world implementation. The observed inconsistencies in surveillance intensity, diagnostic modalities, and follow-up trajectories underscore the need for more standardized, risk-adapted approaches. Notably, the occurrence of symptoms did not appear to influence surveillance patterns, suggesting that surveillance was not strongly guided by patient-reported concerns. These insights can support future efforts to optimize NMIBC surveillance, reduce unwarranted practice variation, and enhance both patient outcomes and the efficient use of healthcare resources.

## Data Availability

The data used for this study are available upon reasonable request. All data requests including NCR data are reviewed by the supervisory committee of the NCR for compliance with the NCR objectives and (inter)national (privacy) regulation and legislation (https://iknl.nl/en/ncr/apply-for-data).

## STATEMENTS

## Acknowledgements

We thank all participants of UroLife and NBCS for their participation. We thank all hospitals involved in UroLife and NBCS. The authors thank the registration team of the Netherlands Comprehensive Cancer Organisation (IKNL) for the collection of data for the Netherlands Cancer Registry. The authors thank Hande Ekmen and Eveline Wijnen from the Department of Research & Development, Research, IKNL, for the collection and quality control of additional data. We thank Dr. Maarten J. Bijlsma from the department of Research & Development, Clinical Data Science, IKNL, for statistical advice. We thank Anja van Gestel from the Department of Research & Development, Clinical Data Science, IKNL, for advice on data visualization.

## Declaration of interests

LH, SV, JH, AG, AV, LK, and KA report no conflicts of interest.

### Funding

The UroLife study was financially supported by Alpe d’HuZes/Dutch Cancer Society (KUN 2013-5926) and Dutch Cancer Society (2017-2/11179). The funding agencies had no further role in this study.

### Author contributions

Lisa M.C. van Hoogstraten: Writing − original draft, Formal analysis, Data curation, Visualization, Conceptualization

Sita H. Vermeulen: Writing − review and editing, Resources

Jasper P. Hof: Writing − review and editing, Data curation

Antoine G. van der Heijden: Writing − review and editing

Alina Vrieling: Writing − review and editing, Resources

Lambertus A. Kiemeney: Writing − review and editing

Katja K.H. Aben: Writing – review and editing, Conceptualization, Project administration

## ONLINE SUPPORTING MATERIAL

**Supplementary Table 1**: Number of cystoscopies per year of follow-up, stratified by risk group

**Supplementary Table 2**: Time interval (in days) between consecutive cystoscopies, stratified by risk group

**Supplementary Table 3**: Patient, tumour and treatment characteristics of a random subset of NMIBCs (primary and recurrent) for whom detailed follow-up information was collected, in total and by risk group

**Supplementary Figure 1**: Swimmer plot depicting individual patient trajectories for follow-up of low risk NMIBC

**Supplementary Figure 2**: Swimmer plot depicting individual patient trajectories for follow-up of intermediate risk NMIBC

**Supplementary Figure 3**: Swimmer plot depicting individual patient trajectories for follow-up of high/very high risk NMIBC

## Notes

### Competing Interest Statement

The authors have declared no competing interest.

### Author Declarations

Both the UroLife study (CMO 2013-494) and NBCS (CMO 2005-315) were approved by an accredited ethics committee in the Netherlands. Ethical approval for the UroLife study was provided by the Committee for Human Research region Arnhem-Nijmegen (CMO 2013-494). All participants provided informed consent. Ethical approval for the NBCS study was provided by the Committee for Human Research region Arnhem-Nijmegen (CMO 2005-315). All participants provided informed consent. In 2021, the name of the Committee for Human Research region Arnhem-Nijmegen changed to Medical Research Ethics Committee (MREC) Oost-Nederland.

