## Supporting material for "Surveillance patterns in non-muscle invasive bladder cancer across risk groups: a real-world analysis"

**Affiliations**

<u>Supporting material</u>

**Supplementary Table 1: Number of cystoscopies per year of follow-up, stratified by risk group**

| Risk group | Year of follow-up | Recommended number of cystoscopies* | N | Mean | SD | Range | Less than recommended (N, %) | | As recommended (N, %) | | More than recommended (N, %) | |
|---|---|---|---|---|---|---|---|---|---|---|---|---|
| **Low risk** | 1 | 1 | 346 | 1.3 | 1.1 | 0-4 | 86 | 24.9 | 130 | 37.6 | 130 | 37.6 |
| | 2 | 1 | 304 | 1.1 | 1.0 | 0-4 | 87 | 28.6 | 119 | 39.1 | 98 | 32.2 |
| | 3 | 1 | 277 | 1.0 | 0.7 | 0-4 | 73 | 26.4 | 138 | 49.8 | 66 | 23.8 |
| | 4 | 1 | 234 | 1.0 | 0.7 | 0-3 | 58 | 24.8 | 133 | 56.8 | 43 | 18.4 |
| | 5 | 1 | 109 | 0.9 | 0.6 | 0-2 | 30 | 27.5 | 65 | 59.6 | 14 | 12.8 |
| **Intermediate risk** (EAU 2016 guideline) | 1 | 1-4 | 665 | 1.7 | 1.2 | 0-5 | 144 | 21.7 | 519 | 78.1 | 2 | 0.3 |
| | 2 | 1-3 | 487 | 1.4 | 1.1 | 0-6 | 137 | 28.1 | 342 | 70.2 | 8 | 1.6 |
| | 3 | 1-2 | 395 | 1.1 | 0.9 | 0-4 | 123 | 31.1 | 246 | 62.3 | 26 | 6.6 |
| | 4 | 1-2 | 283 | 0.9 | 0.8 | 0-3 | 100 | 35.3 | 176 | 62.2 | 7 | 2.5 |
| | 5 | 1-2 | 108 | 0.8 | 0.7 | 0-3 | 43 | 39.8 | 64 | 59.3 | 1 | 0.9 |
| **Intermediate risk** (EAU 2024 guideline) | 1 | 2 | 665 | 1.7 | 1.2 | 0-5 | 292 | 43.9 | 182 | 27.4 | 191 | 28.7 |
| | 2 | 2 | 487 | 1.4 | 1.1 | 0-6 | 256 | 52.6 | 147 | 30.2 | 84 | 17.3 |
| | 3 | 1 | 395 | 1.1 | 0.9 | 0-4 | 123 | 31.1 | 146 | 37.0 | 126 | 31.9 |
| | 4 | 1 | 283 | 0.9 | 0.8 | 0-3 | 100 | 35.3 | 126 | 44.5 | 57 | 20.1 |
| | 5 | 1 | 108 | 0.8 | 0.7 | 0-3 | 43 | 39.8 | 50 | 46.3 | 15 | 13.9 |
| **High/very high risk** | 1 | 4 | 889 | 2.3 | 1.2 | 0-5 | 784 | 88.2 | 101 | 11.4 | 4 | 0.5 |
| | 2 | 3 | 665 | 2.2 | 1.3 | 0-5 | 353 | 53.1 | 206 | 31.0 | 106 | 15.9 |
| | 3 | 2 | 520 | 1.6 | 1.2 | 0-5 | 202 | 38.9 | 208 | 40.0 | 110 | 21.2 |
| | 4 | 2 | 387 | 1.3 | 1.0 | 0-4 | 201 | 51.9 | 163 | 42.1 | 23 | 5.9 |
| | 5 | 2 | 175 | 1.0 | 0.8 | 0-3 | 122 | 69.7 | 49 | 28.0 | 4 | 2.3 |

* The average number of cystoscopies was calculated for each consecutive year <u>after</u> the first surveillance cystoscopy. For example, for HR NMIBC, cystoscopy is recommended every 3 months for 2 years, i.e., 4 cystoscopies per year, and then every 6 months up to 5 years, i.e., 2 cystoscopies per year. Omitting the first surveillance cystoscopy results in 4 recommended cystoscopies in the first year, and 3 cystoscopies in the second year.

SD: standard deviation

**Supplementary Table 2: Time interval (in days) between consecutive cystoscopies, stratified by risk group**

| Risk group | Cystoscopy | N | Time interval as recommended* (days) | Mean time interval | SD | Difference | Median time interval | IQR | Difference |
|---|---|---|---|---|---|---|---|---|---|
| Low risk | 1 and 2 | 312 | 270 | 160.7 | 82.3 | -109.3 | 138.5 | 98.0 - 188.5 | -131.5 |
| | 2 and 3 | 270 | 365 | 219.8 | 121.8 | -145.2 | 188.5 | 114.0 - 289.0 | -176.5 |
| | 3 and 4 | 249 | 365 | 250.2 | 139.9 | -114.8 | 196.0 | 127.0 - 369.0 | -169 |
| | 4 and 5 | 221 | 365 | 271.4 | 115.7 | -93.6 | 280.0 | 178.0 - 371.0 | -85 |
| | 5 and 6 | 201 | 365 | 280.0 | 121.2 | -85.0 | 336.0 | 182.0 - 371.0 | -29 |
| | 6 and 7 | 154 | | 270.9 | 114.3 | | 279.5 | 175.0 - 366.0 | |
| | 7 and 8 | 101 | | 246.9 | 137.3 | | 197.0 | 169.0 - 363.0 | |
| | 8 and 9 | 79 | | 268.3 | 172.2 | | 197.0 | 183.0 - 367.0 | |
| | 9 and 10 | 56 | | 242.9 | 125.0 | | 192.0 | 175.0 - 333.0 | |
| | 10 and 11 | 41 | | 224.2 | 97.2 | | 191.0 | 179.0 - 287.0 | |
| | 11 and 12 | 30 | | 288.0 | 155.7 | | 200.0 | 182.0 - 367.0 | |
| | 12 and 13 | 13 | | 237.5 | 106.4 | | 190.0 | 168.0 - 350.0 | |
| | 13 and 14 | 10 | | 275.9 | 102.3 | | 275.5 | 182.0 - 372.0 | |
| | 14 and 15 | 5 | | 292.0 | 105.7 | | 364.0 | 183.0 - 364.0 | |
| | 15 and 16 | 1 | | 351.0 | | | 351.0 | 351.0 - 351.0 | |
| Intermediate risk* | 1 and 2 | 718 | 180 | 144.3 | 86.5 | -35.7 | 119.0 | 93.0 - 182.0 | -61.0 |
| | 2 and 3 | 593 | 180 | 164.6 | 112.8 | -15.4 | 126.0 | 98.0 - 189.0 | -54.0 |
| | 3 and 4 | 494 | 180 | 184.6 | 106.9 | 4.6 | 154.0 | 98.0 - 215.0 | -26.0 |
| | 4 and 5 | 410 | 180 | 198.6 | 104.5 | 18.6 | 181.0 | 119.0 - 238.0 | 1.0 |
| | 5 and 6 | 337 | 360 | 212.6 | 108.7 | -147.4 | 183.0 | 124.0 - 280.0 | -182.3 |
| | 6 and 7 | 264 | 360 | 221.4 | 110.1 | -138.6 | 188.0 | 133.0 - 297.0 | -177.3 |
| | 7 and 8 | 198 | 360 | 211.9 | 104.8 | -148.1 | 187.5 | 146.0 - 260.0 | -177.8 |
| | 8 and 9 | 149 | 360 | 213.7 | 114.3 | -146.3 | 188.0 | 147.0 - 253.0 | -177.3 |
| | 9 and 10 | 111 | 360 | 239.0 | 114.9 | -121.0 | 203.0 | 162.0 - 364.0 | -162.3 |
| | 10 and 11 | 78 | 360 | 228.0 | 101.9 | -132.0 | 196.0 | 172.0 - 353.0 | -169.3 |
| | 11 and 12 | 41 | | 186.7 | 84.7 | | 178.0 | 125.0 - 219.0 | |
| | 12 and 13 | 23 | | 191.5 | 97.8 | | 186.0 | 142.0 - 203.0 | |
| | 13 and 14 | 13 | | 225.8 | 124.1 | | 192.0 | 134.0 - 377.0 | |
| | 14 and 15 | 9 | | 226.2 | 73.7 | | 190.0 | 181.0 - 252.0 | |
| | 15 and 16 | 5 | | 273.2 | 95.7 | | 276.0 | 184.0 - 365.0 | |
| | 16 and 17 | 3 | | 244.0 | 147.3 | | 277.0 | 83.0 - 372.0 | |
| High/very high risk | 1 and 2 | 993 | 90 | 117.2 | 75.6 | 27.2 | 99.0 | 91.0 - 124.0 | 9.0 |
| | 2 and 3 | 881 | 90 | 115.3 | 48.9 | 25.3 | 98.0 | 91.0 - 126.0 | 8.0 |
| | 3 and 4 | 785 | 90 | 119.1 | 51.7 | 29.1 | 98.0 | 91.0 - 129.0 | 8.0 |
| | 4 and 5 | 683 | 90 | 127.8 | 59.1 | 37.8 | 105.0 | 92.0 - 148.0 | 15.0 |
| | 5 and 6 | 623 | 90 | 138.8 | 69.8 | 48.8 | 117.0 | 95.0 - 181.0 | 27.0 |
| | 6 and 7 | 545 | 90 | 153.2 | 83.6 | 63.2 | 126.0 | 97.0 - 187.0 | 36.0 |
| | 7 and 8 | 466 | 90 | 162.0 | 77.4 | 72.0 | 148.0 | 99.0 - 190.0 | 58.0 |
| | 8 and 9 | 396 | 180 | 168.8 | 74.7 | -11.2 | 169.0 | 109.5 - 195.0 | -11.0 |
| | 9 and 10 | 328 | 180 | 182.0 | 83.4 | 2.0 | 182.0 | 118.5 - 203.0 | 2.0 |
| | 10 and 11 | 264 | 180 | 181.3 | 79.6 | 1.3 | 183.0 | 121.5 - 200.0 | 3.0 |
| | 11 and 12 | 206 | 180 | 189.2 | 78.4 | 9.2 | 186.0 | 140.0 - 203.0 | 6.0 |
| | 12 and 13 | 154 | 180 | 202.7 | 89.6 | 22.7 | 189.0 | 145.0 - 237.0 | 9.0 |
| | 13 and 14 | 103 | 180 | 225.0 | 101.4 | 45.0 | 189.0 | 165.0 - 315.0 | 9.0 |
| | 14 and 15 | 60 | 180 | 215.8 | 103.9 | 35.8 | 182.5 | 143.5 - 307.5 | 2.5 |
| | 15 and 16 | 33 | 180 | 204.5 | 94.3 | 24.5 | 190.0 | 140.0 - 224.0 | 10.0 |
| | 16 and 17 | 19 | 180 | 229.9 | 104.1 | 49.9 | 182.0 | 155.0 - 364.0 | 2.0 |

* For the intermediate risk group, the difference was calculated for the EAU 2024 guidelines. Earlier guidelines, using an in-between individualized scheme, do not mention specific time intervals.

SD: standard deviation; IQR: interquartile range

**Supplementary Table 3: Patient, tumour and treatment characteristics of a random subset of NMIBCs (primary and recurrent) for whom detailed follow-up information was collected, in total and by risk group**

| | All NMIBCs (N=204) | | Low risk (N=44) | | Intermediate risk (N=88) | | High/very high risk (N=72) | |
|---|---|---|---|---|---|---|---|---|
| | N | (%) | N | (%) | N | (%) | N | (%) |
| Patient characteristics | | | | | | | | |
| **Age at diagnosis** (median, IQR) | 65.5 | (59.0-72.0) | 65.5 | (56.0-71.0) | 61.5 | (54.0-72.5) | 67.5 | (62.0-73.0) |
| **Gender** (% female) | 48 | (23.5%) | 14 | (31.8%) | 18 | (20.5%) | 16 | (22.2%) |
| **Smoking status** | | | | | | | | |
| Never smoker | 46 | (22.5%) | 8 | (18.2%) | 23 | (26.1%) | 15 | (20.8%) |
| Former smoker | 107 | (52.5%) | 21 | (47.7%) | 51 | (58.0%) | 35 | (48.6%) |
| Current smoker | 48 | (23.5%) | 14 | (31.8%) | 14 | (15.9%) | 20 | (27.8%) |
| Unknown | 3 | (1.5%) | 1 | (2.3%) | - | | 2 | (2.8%) |
| **Hospital of diagnosis** | | | | | | | | |
| Teaching hospital | 145 | (71.1%) | 28 | (63.6%) | 65 | (73.9%) | 52 | (72.2%) |
| Academic hospital | 59 | (28.9%) | 16 | (36.4%) | 23 | (26.1%) | 20 | (27.8%) |
| **Hospital ID** | | | | | | | | |
| Hospital 1 | 8 | (3.9%) | 8 | (18.2%) | - | | - | |
| Hospital 2 | 67 | (32.8%) | 5 | (11.4%) | 33 | (37.5%) | 29 | (40.3%) |
| Hospital 3 | 59 | (28.9%) | 16 | (36.4%) | 23 | (26.1%) | 20 | (27.8%) |
| Hospital 4 | 70 | (34.3%) | 15 | (34.1%) | 32 | (36.4%) | 23 | (31.9%) |
| Tumour characteristics | | | | | | | | |
| **Clinical tumour stage** | | | | | | | | |
| Ta | 146 | (71.6%) | 44 | (100.0%) | 88 | (100.0%) | 14 | (19.4%) |
| T1 | 38 | (18.6%) | - | | - | | 38 | (52.8%) |
| CIS (Tai, Tis, T1i) | 20 | (9.8%) | - | | - | | 20 | (27.8%) |
| **Grade of tumour (low/high)** | | | | | | | | |
| Low grade (G1, G2, G2A) | 146 | (71.6%) | 44 | (100.0%) | 88 | (100.0%) | 14 | (19.4%) |
| High grade (G2B, G3) | 56 | (27.5%) | - | | - | | 56 | (77.8%) |
| Unknown | 2 | (1.0%) | - | | - | | 2 | (2.8%) |
| **Focality of the tumour** | | | | | | | | |
| Unifocal | 149 | (73.0%) | 44 | (100.0%) | 57 | (64.8%) | 48 | (66.7%) |
| Multifocal | 54 | (26.5%) | - | | 31 | (35.2%) | 23 | (31.9%) |
| Unknown | 1 | (0.5%) | - | | - | | 1 | (1.4%) |
| **Primary or recurrent tumour** | | | | | | | | |
| Primary | 159 | (77.9%) | 44 | (100.0%) | 56 | (63.6%) | 59 | (81.9%) |
| Recurrent | 45 | (22.1%) | - | | 32 | (36.4%) | 13 | (18.1%) |
| Treatment characteristics | | | | | | | | |
| **reTURBT** | 57 | (27.9%) | 5 | (11.4%) | 10 | (11.4%) | 42 | (58.3%) |
| **Chemotherapy** | 139 | (68.1%) | 35 | (79.5%) | 69 | (78.4%) | 35 | (48.6%) |
| **BCG** | 58 | (28.4%) | - | | 6 | (6.8%) | 52 | (72.2%) |
| Follow-up | | | | | | | | |
| **At least 1 cystoscopy** | 199 | (97.5%) | 44 | (100.0%) | 85 | (96.6%) | 70 | (97.2%) |
| **At least 1 biopsy** | 99 | (48.5%) | 11 | (25.0%) | 32 | (36.4%) | 56 | (77.8%) |
| **At least 1 CT urography/IVP** | 76 | (37.3%) | 11 | (25.0%) | 25 | (28.4%) | 40 | (55.6%) |
| **At least 1 cytology** | 132 | (64.7%) | 22 | (50.0%) | 46 | (52.3%) | 64 | (88.9%) |
| **At least 1 symptom (incl. haematuria)** | 68 | (33.3%) | 20 | (45.5%) | 23 | (26.1%) | 25 | (34.7%) |

NMIBC: non-muscle invasive bladder cancer; IQR: interquartile range; TURBT: transurethral resection of the bladder tumour; BCG: Bacillus Calmette-Guérin; CT: computed tomography; IVP: intravenous pyelogram

**Supplementary Figure 1: Swimmer plot depicting individual patient trajectories for follow-up of low risk NMIBC**

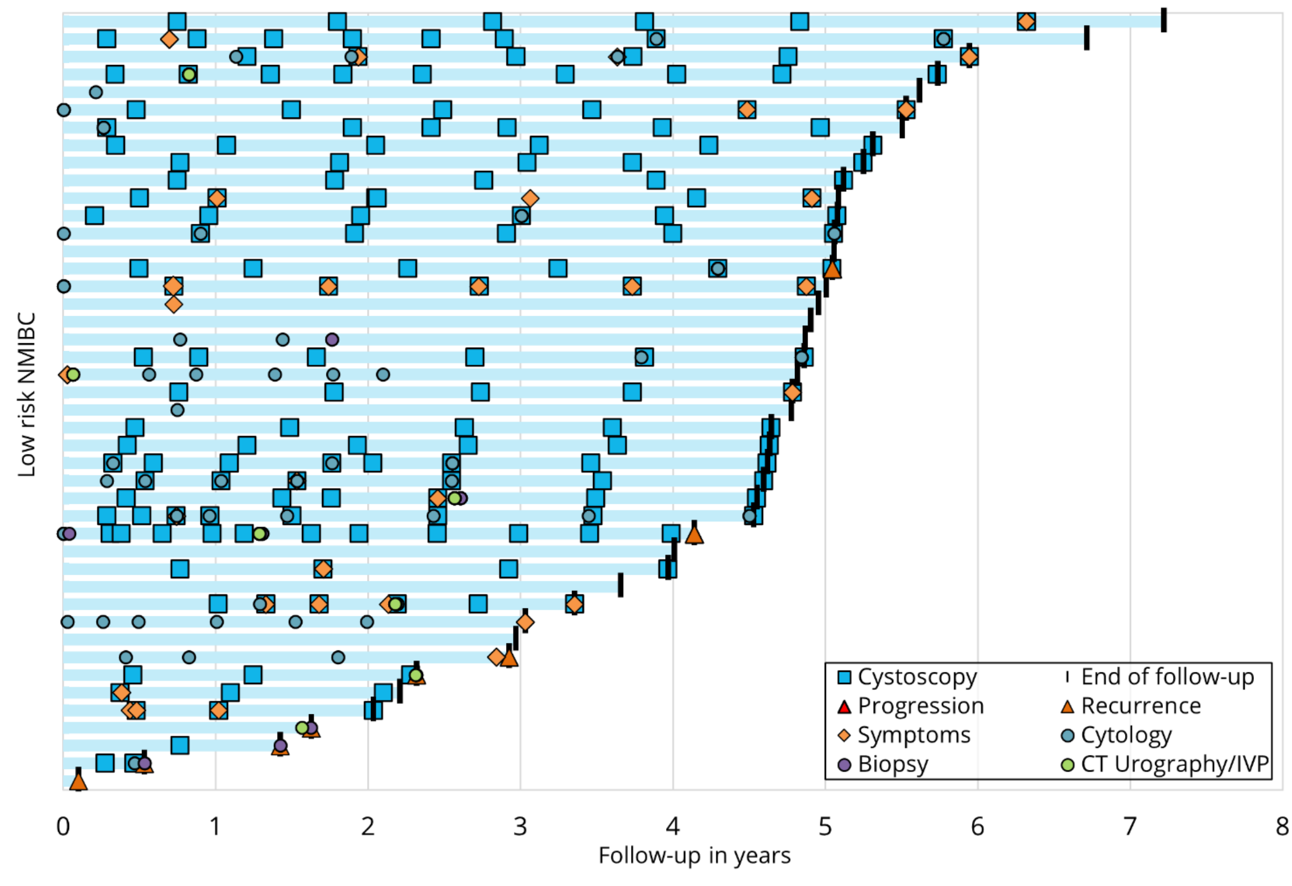


NMIBC: non-muscle invasive bladder cancer; CT: computed tomography; IVP: intravenous pyelogram

**Supplementary Figure 2: Swimmer plot depicting individual patient trajectories for follow-up of intermediate risk NMIBC**

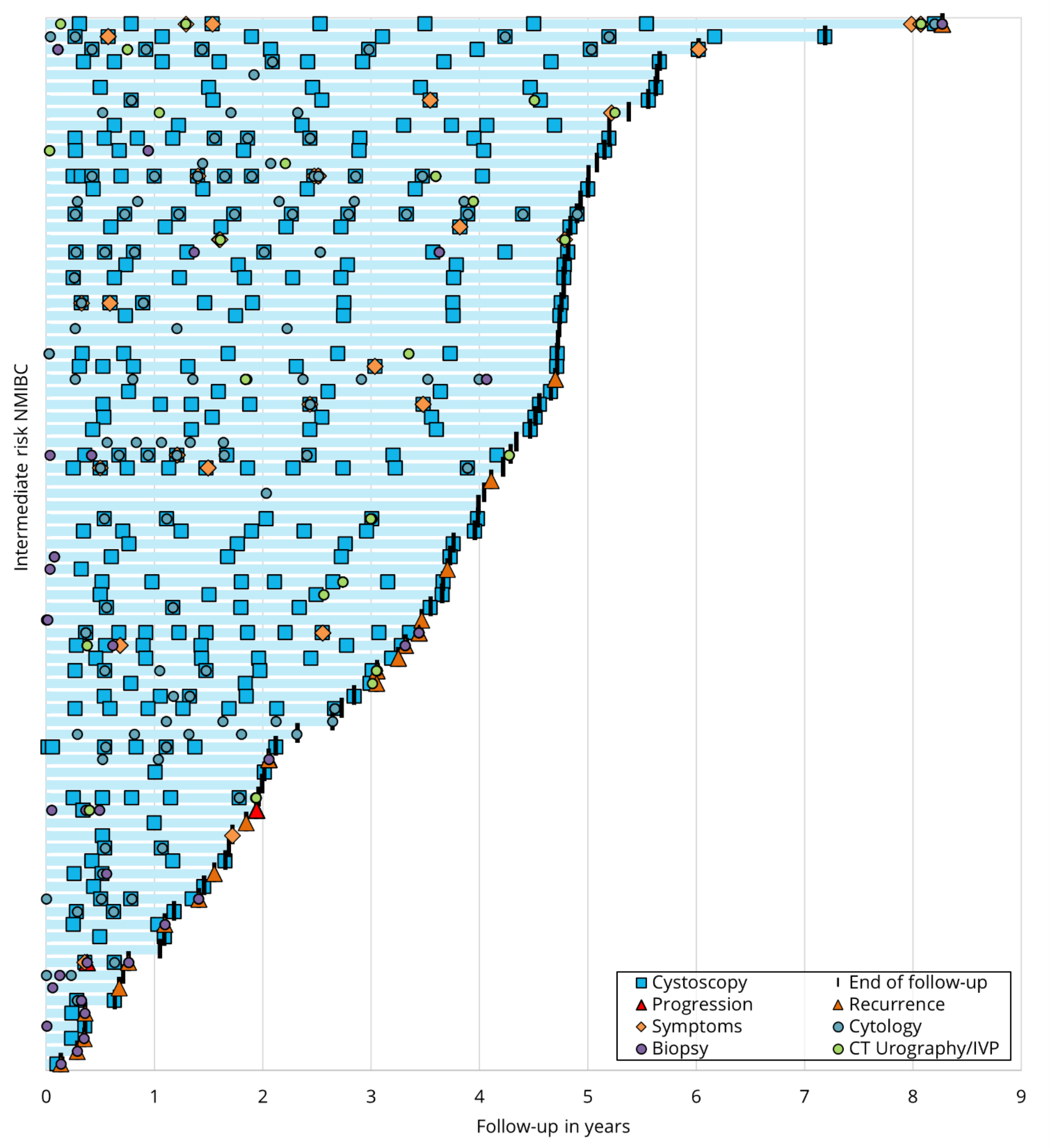


NMIBC: non-muscle invasive bladder cancer; CT: computed tomography; IVP: intravenous pyelogram

**Supplementary Figure 3: Swimmer plot depicting individual patient trajectories for follow-up of high/very high risk NMIBC**

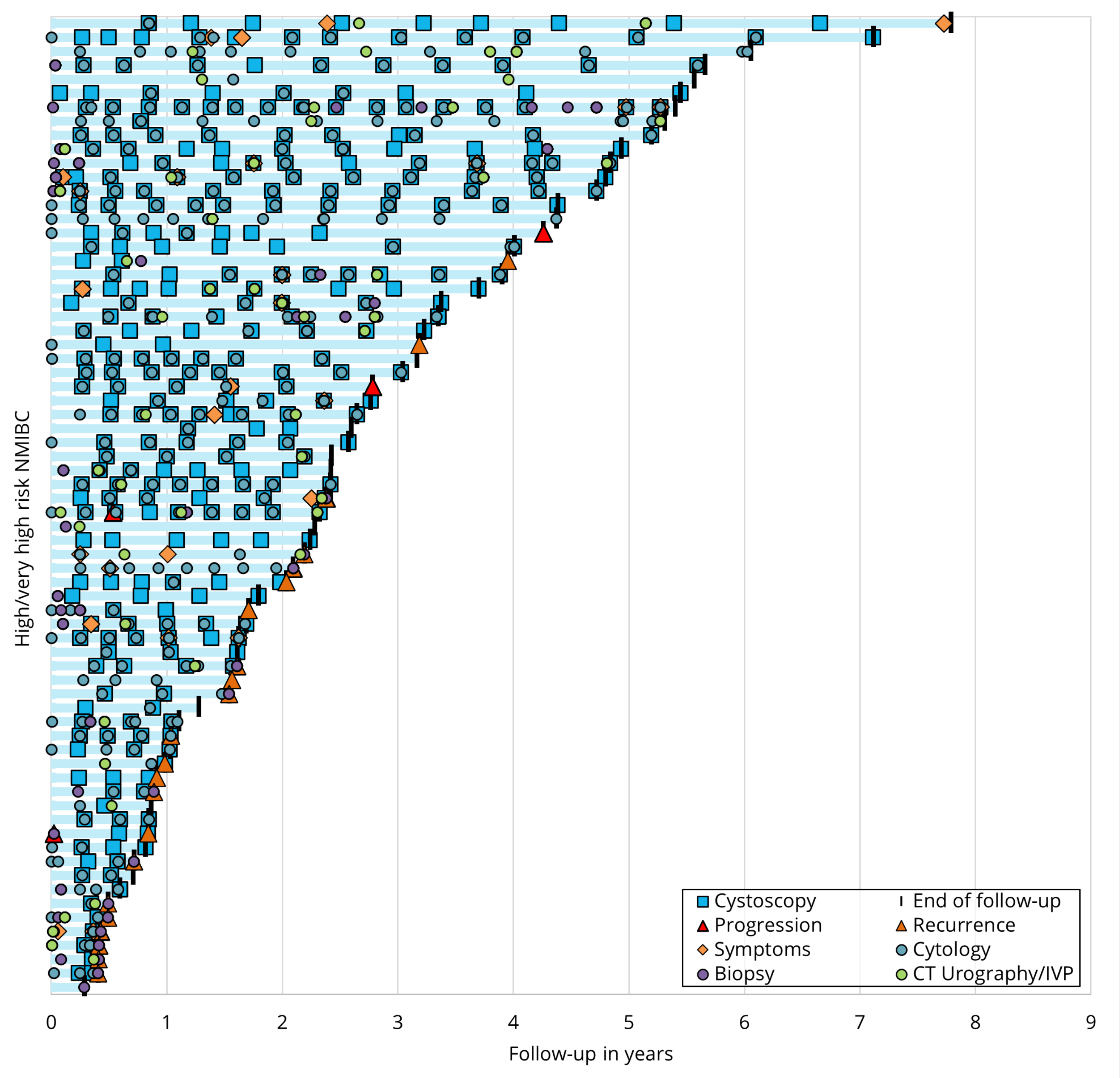


NMIBC: non-muscle invasive bladder cancer; CT: computed tomography; IVP: intravenous pyelogram
